# *Anopheles* Mosquito Distribution and Malaria Outbreak Risk Assessment in Georgia, USA (2001–2025): Implications for Surveillance and Preparedness

**DOI:** 10.64898/2026.10.22.26366233

**Authors:** Bridget O. Aito-Bobadoye, Adem Filali, Elmer W. Gray, Skyler M. Kerr, Tiffany Nguyen, Rosemary Kelly

## Abstract

Competent malaria vector species in Georgia represents an emerging public health threat. Despite the successful elimination of endemic malaria transmission from the United States, the entomological conditions necessary for re-establishment of local transmission still persists across ecologically permissive regions of the south-eastern United States. However, no comprehensive, county-level assessment of *Anopheles* mosquito distribution and associated malaria outbreak risk has been conducted across the state. This study analyzed 2,716 *Anopheles* collection records, representing 11,265 individual specimens, from 24 years of surveillance data (2001–2025). Results classified Fulton County as a high risk hot spot and critical convergence zone for malaria outbreak with the most significantly highest risk score (25), *Anopheles* population density (2,561) and elevated MIR activity (> 4.1%). Glynn and Lowndes counties were designated as moderate risk (15 and 12 respectively), driven primarily by substantial *Anopheles* populations. Conversely, DeKalb County, despite exhibiting the highest individual infection rate (MIR: 6.67%), was classified low risk (Score: 10) due to lower *Anopheles* abundance. Our results underscore the need for improved mosquito surveillance in high-risk counties, to address the dual health threat of arboviral and potential human malaria outbreaks under current climate change scenarios in Georgia.

## Introduction

Malaria is one of the most debilitating infectious diseases afflicting human populations, been one of the most prevalent and globally widespread parasitic diseases. It is caused by obligate intracellular protozoan parasites of the genus *Plasmodium* (Plasmodiidae, Haemosporida). This parasite is responsible for the acute febrile syndrome transmitted to humans through the bite of infected female *Anopheles* mosquitoes (Diptera : Culicidae) (1). Five *Plasmodium* species have been identified as etiological agents of human malaria : *P. vivax, P. falciparum, P. malariae*, P. *ovale*, and *P. knowlesi* (1, 2). Among these, both *P. falciparum* and *P. vivax*, account for the greatest burden of human infections globally (2). Nonetheless, the most clinically severe and potentially fatal form of malaria is attributable to *P. falciparum*, which is predominantly distributed across the tropical regions of the world especially Asia and Sub-Saharan Africa (2). Children under five years of age and pregnant women represent the most vulnerable demographic groups (1, 3, 4).

The epidemiology of malaria is geographically defined, especially for people residing in or traveling to sub-Saharan Africa, Southeast Asia, the Eastern Mediterranean, the Western Pacific, and the Americas are highly vulnerable and at considerable risk of infection (2, 3). The burden of malaria is disproportionately concentrated in sub-Saharan Africa, which account for approximately 90% of malaria cases and 94% of all global malaria-related deaths world-wide (3,4). Malaria was historically endemic in the southeastern United States until its elimination in the 1950s through conceited interventions such as improved sanitation infrastructure, and widespread application of insecticides and executed by the Office of Malaria Control in War Areas, currently known as the Center for Disease Control and Prevention (CDC) (1,2, 5,9). The principal historical vectors of malaria are *Anopheles quadrimaculatus* in the eastern United States and *An. freeborni* Aitken in the western United States. Additional competent vectors include *An. punctipennis* (Say), *An. albimanus Wiedemann, An. pseudopunctipennis* Theobald, members of the *An. quadrimaculatus* complex (*An. quadrimaculatus, An. diluvialis, An. inundatus, An. maverlius, and An. smaragdinus*), and members of the *An. crucians Wiedemann* complex (*An. crucians, An. bradleyi*, and *An. georgianus*) (1). Despite the complete elimination of endemic transmission, sporadic cases of locally acquired malaria still continue to be reported across the United States (10, 12).

In 2023 confirmed cases of autochthonous malaria were detected in Florida and Texas respectively. This was the first report of locally transmitted cases in the United States in over two decades and has renewed valid concerns about the potential re-emergence of malaria transmission in several parts of southeastern United States. Georgia, sharing similar ecological and climatic characteristics with Florida, represents a high-priority threat for renewed surges in human malaria cases as depicted from ongoing mosquito surveillance across the state.

The state of Georgia has recorded multiple *Anopheles* species confirmed to be historically associated with malaria transmission, including *Anopheles quadrimaculatus*, the primary historical malaria vector in the eastern United States, *Anopheles punctipennis*, a confirmed vector of *Plasmodium vivax*, and *Anopheles crucians*, another mosquito species associated with malaria transmission in southeastern wetland ecosystems (6,7). Despite their documented presence, systematic surveillance of these *Anopheles* species remains fragmented, creating significant gaps in understanding their bionomics especially their current distribution, life-history traits, behaviors such as biting behavior, main and alternative hosts for blood meals and finally ecological niches or possible co-occurrence with other mosquito populations in Georgia.

Available epidemiological reports across the United States indicate a sustained upward trend in the incidence of malaria cases over recent years. Surveillance data compiled from the period spanning 2011 to 2016 revealed an annual mean of 1,773 malaria cases reported across the country (4,10-12, 18–21). 2016 marked a significant epidemiological milestone, with records confirming a total of 2,078 cases, the highest annual count documented in the United States since 1972 (4). Further analysis of the geographic distribution of malaria burden in the United States revealed that ten states disproportionately accounted for 64.1% of all reported cases, with respective annual mean case counts obtained across the following states: New York (296.4), Maryland (147.4), California (118.2), Texas (115.6), New Jersey (91.6), Georgia (77.6), Virginia (77.4), Florida (77.0), Massachusetts (75.2), and Pennsylvania (73.6) (4,17, 18–21).

Epidemiological data from verified sources indicate that approximately 98.8% of malaria cases reported in the United States are possibly of imported or acquired origin, with the majority (75.0%) attributed to individuals returning from sub-Saharan African nations (4). With respect to plasmodium species distribution, *P. falciparum* and *P. vivax* constitute the two predominant species identified among returning American travelers, accounting for 61.9% and 15.1% of confirmed cases, respectively (4, 15,18–21). Notably, only one single importation of *P. knowles*i has been documented in the United States to date, and this case was identified in a traveler returning from the Philippines (22, 23, 24, 25, 26). During the surveillance period spanning 2011 to 2016, no autochthonous malaria transmission was recorded in the United States (4).

A principal public Heath threat in the context of malaria in the state of Georgia pertains to the potential re-emergence and establishment of endemic transmission, not withstanding the considerably reduced likelihood of sustained local transmissions. Several deterministic factors have been identified as pre-requisites for the re-establishment of endemic malaria transmission, including, but not limited to the presence of competent *Anopheles* vectors, the availability of suitable climatic and environmental conditions necessary for the development of *Anopheles* mosquitoes and the survival of plasmodium parasite (26, 27). The preponderance of contemporary literature addressing malaria in the United States remains largely concentrated on therapeutic regimens and the economic burden associated with treatment, rather than on transmission risk assessment of competent mosquito vectors (28–30). The objective of this study was to assess the population distribution of *Anopheles* mosquito species and identify both high risk hot-spot and critical convergence zones of possible malaria out breaks in the state of Georgia.

In 2003, epidemiological reports from the CDC reported four instances of locally acquired, endemic malaria transmission, with the most recently reported outbreak occurring in Palm Beach County, Florida. Entomological surveillance conducted during the course of this period failed to yield any Plasmodium-positive female *Anopheles* specimens, nevertheless, *An. quadrimaculatus* and *An. crucians* have been designated as the most plausible mosquito vectors on the basis of their established biological competence and geographic distribution (17). Across all recorded out breaks, *P. vivax* was consistently identified as the implicated plasmodium species, with epidemiological evidence highlighting migrant laborers of Mexican origin as the probable reservoir of parasite introduction into Florida (17).

The two-decade absence of confirmed endemic malaria transmission following the confirmed 2003 outbreak does not, however, diminish the imperative for continued mosquito surveillance. Ecological, climatic, and demographic conditions conducive for localized transmission still remain fully functional across multiple states in the country, which necessitates improvement in surveillance infrastructure and response capacities at both the local and state levels. It is within this framework that the present study advances county-level risk assessment of plausible malaria outbreak by estimating *Anopheles* distribution across the state of Georgia. By understanding the population dynamics of *Anopheles* distribution, delineating high-risk transmission convergence zones, and situating these findings within a broader trajectory of climate-driven ecological transmissions, this study provides a critical evidence basis for improved but targeted surveillance of *Anopheles* mosquitoes.

## Materials and Methods

### Study Area and Geographical Scope

Mosquito surveillance has been conducted across the state of Georgia from 2001-2025, which encompasses all counties distributed across six distinct ecological geo-regions: the Blue Ridge, Piedmont, Coastal Plain, and Valley and Ridge (Figure 1). These geo-regions exhibit considerable heterogeneity in land use, hydrology, and climate, which provides an ecologically diverse landscape for mosquito breeding. Georgia’s subtropical humid climate, characterized by warm temperatures, high annual precipitation, and extensive wetland ecosystems, renders the state particularly conducive to the establishment of *Anopheles* populations.

**Figure 1.**
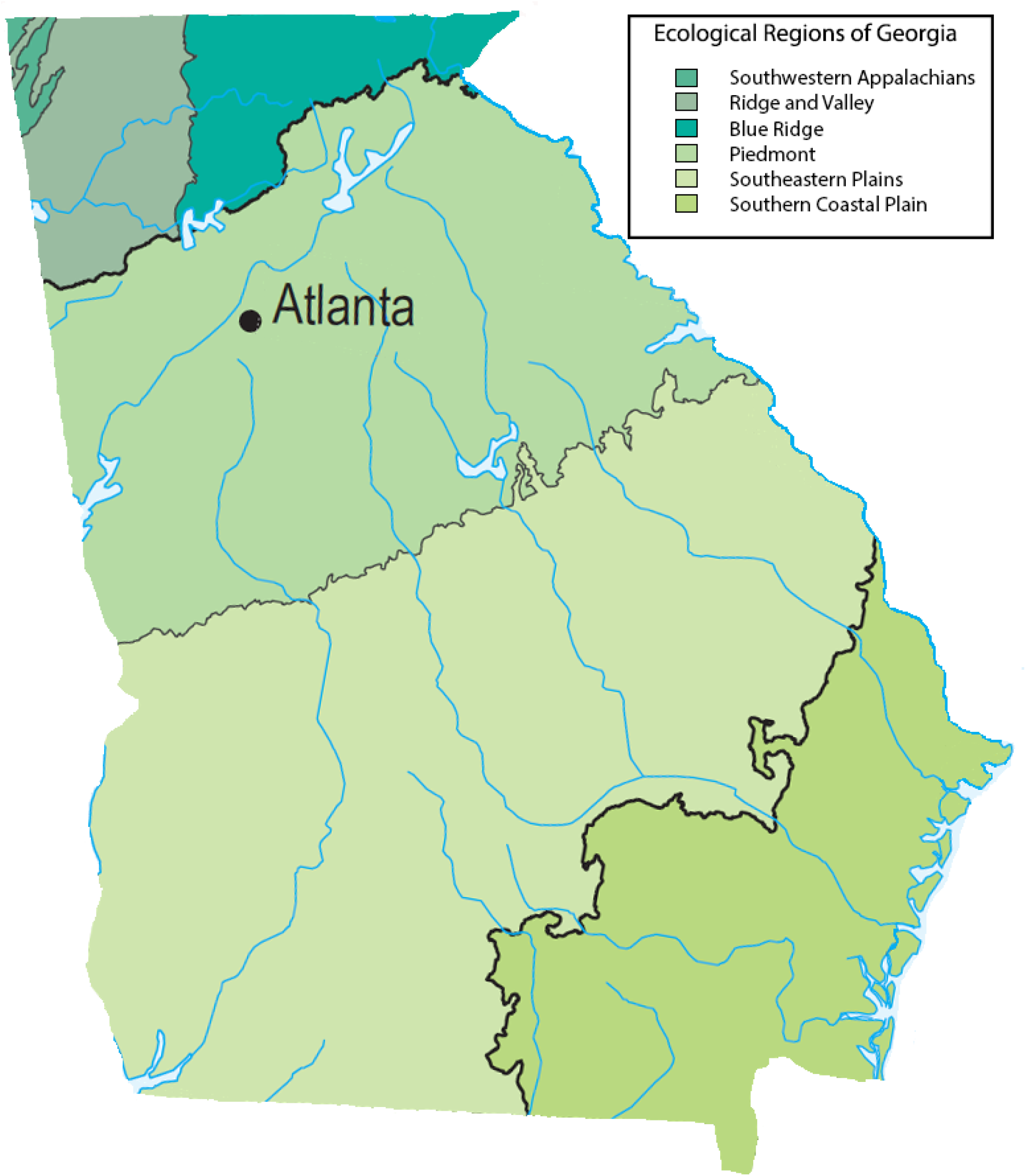
Ecological regions of the state of Georgia.

### Specimen Collection and Species Identification

Adult *Anopheles* mosquito specimens were reported to have been collected using standardized entomological surveillance trapping methods employed by the GDPH surveillance program across all counties. Morphological identification of all collected specimens to species level was performed by trained and experienced entomologists at the GDPH entomological laboratory, by utilizing established dichotomous taxonomic keys and morphological reference criteria.

### Surveillance Data Sources

Entomological surveillance data underpinning this assessment were systematically extracted from the Georgia Department of Public Health (GDPH) mosquito surveillance data base, spanning a continuous 24-year surveillance period from 2001 to 2025. The data-set comprised a total of 11, 271 *Anopheles* specimen records collected from ten Georgia counties where mosquito surveillance is currently ongoing. These inclusive datasets, represent one of the most extensive statewide mosquito surveillance data sets assembled to date from the southeastern region of the United States. It is noted that no *Anopheles* specimen records were detected for the calendar year 2021; this absence is attributed to substantially reduced surveillance activity during the COVID-19 pandemic recovery period rather than a genuine biological absence of vector populations, and the year 2021 was accordingly excluded from temporal trend analyses.

### *Anopheles* Species Abundance and County-Level Distribution Analysis

Quantitative assessment of *Anopheles* species composition was conducted by calculating the total abundance and relative proportional representation of each identified *Anopheles* mosquito species across the full 24-year surveillance period. Abundance metrics were aggregated at the county level, while annual temporal trends in species-specific *Anopheles* abundance were further examined to elucidate patterns in population dynamics across the study period (2001-2025), with particular attention to the three epidemiologically significant primary malaria vector species : *An. crucians, An. punctipennis*, and *An. quadrimaculatus*. County level *Anopheles* vector abundance was further integrated with WNV Minimum Infection Rate MIR data to identify counties of convergent vector borne disease risk. A fixed threshold quadrant classification was applied based on biologically meaningful cutoff values. Counties were then classified as critical convergence zones when WNV MIR exceeded 1% per 100 mosquito pools and *Anopheles* abundance exceeded 500 specimens over the 24 year study period. Counties meeting only the *Anopheles* threshold were classified as high *Anopheles* risk areas, while counties meeting only the WNV threshold were classified as high WNV risk, the remaining counties were classified as lower risk. All counties with documented *Anopheles* surveillance records were retained regardless of WNV detection status. All analyses were performed using Python software V.3.12.

To conduct a malaria outbreak risk assessment in Georgia under projected future climate change scenarios, we conducted a risk matrix scoring estimation using the Strategic Tool for Assessing Risks (STAR) which was developed by the World Health Organization (WHO). Based on integrated likelihood parameters to classify possible health hazards (malaria) into five risk categories ranging from very low to very high, we combined population level and county-level *Anopheles* vector abundance data derived from existing GDPH surveillance data-sets from 2001-2025 to achieve malaria outbreak risk matrix scores for all surveilled counties. This helped delineate counties exhibiting disproportionately elevated *Anopheles* population densities.

### Statistical Analysis

All statistical analyses were performed using R statistical software (31). Descriptive statistics were used to summarize *Anopheles* species composition, including total specimen counts and proportional abundance for each identified *Anopheles* species from all ten counties across the 24-year surveillance period. Temporal trends in annual *Anopheles* abundance were examined using linear regression, with calendar year as the independent variable and annual specimen count as the outcome.

## Results

### Anopheles Species Composition and Abundance

A total of 2,716 *Anopheles* collection records, representing 11,265 individual specimens, were recorded across Georgia between 2001-2025, representing six species. Three species of primary malaria vector significance dominated all specimen collections, totaling 10,342 specimens: *An. crucians* (5,390 specimens, 52.7%), *An. punctipennis* (3,321 specimens, 28.6%), and *An. quadrimaculatus* (1,631 specimens, 20.2%) (Figure 2). Minor *Anopheles* species including *An. perplexens, An. barberi*, and *An. atropos* represented less than 0.1% of total specimen collections. Surveillance data were continuous across all years except 2021, where no *Anopheles* records were detected, likely reflecting reduced surveillance effort during the COVID-19 pandemic recovery period rather than true absence.

**Figure 2.**
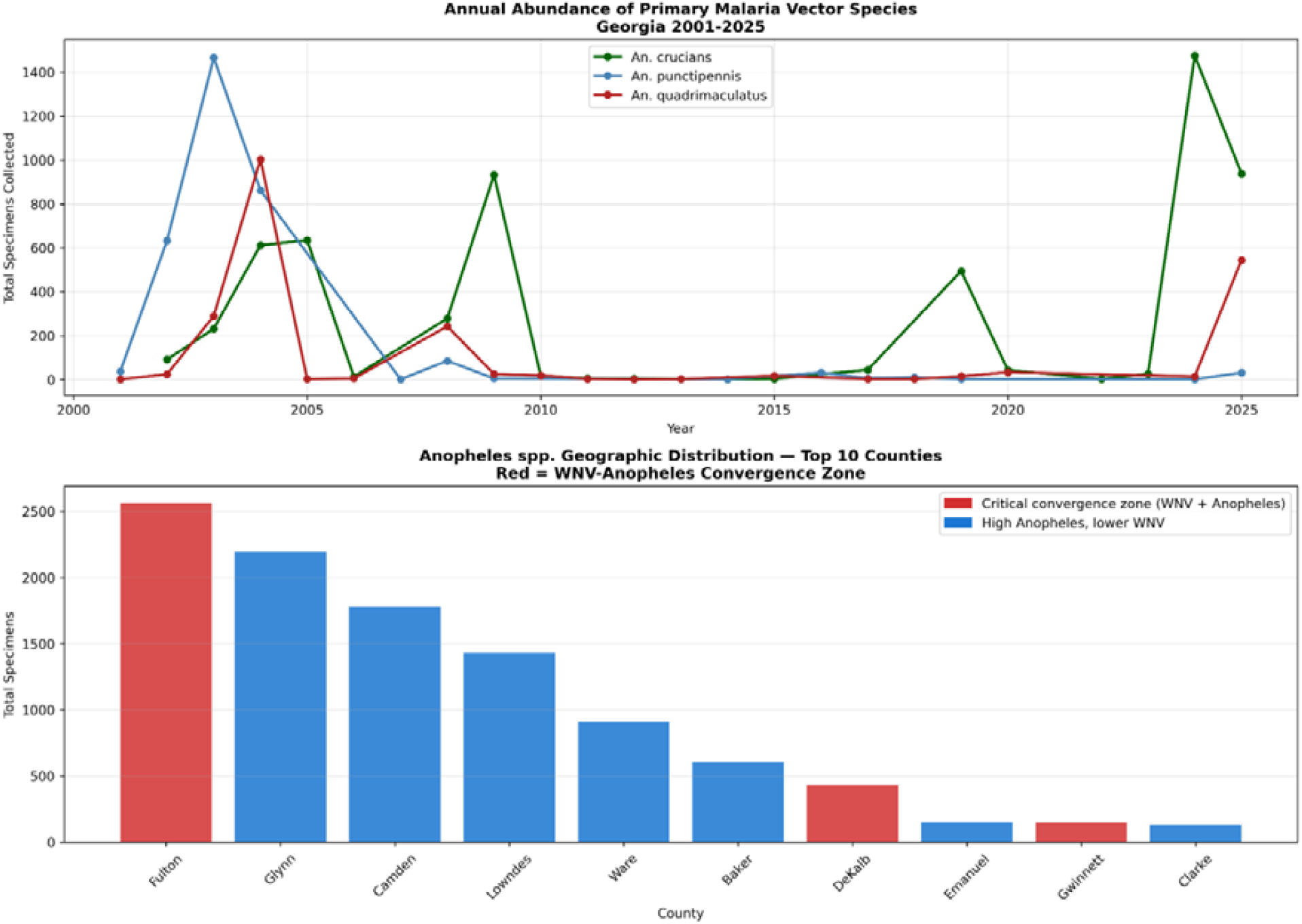
Temporal trends and geographic distribution of *Anopheles* mosquitoes in Georgia (2001– 2025). Panel A: Annual abundance of *An. crucians, An. punctipennis* and *An. quadrimaculatus*. Panel B: Total *Anopheles* specimens by county, highlighting critical WNV-*Anopheles* convergence zones (red) versus high *Anopheles* risk zones (blue).

### Geographic Distribution of *Anopheles* mosquitoes

*Anopheles* abundance was highest in Fulton County (2,561 specimens), followed by Glynn (2,193), Camden (1,780), Lowndes (1,430), and Ware (911) counties (Table 1; Figure 2). *Anopheles* mosquito abundance identified Fulton County as the most critical convergence zone and a confirmed likelihood hot spot for future malaria outbreaks under current climate change scenarios.

**Table 1:** Malaria outbreak risk scores for counties with recorded *Anopheles* mosquito species.

| County | *Anopheles*<br>Specimens | WNV MIR (%) | STAR Likelihood | STAR Impact | Risk Score |
| --- | --- | --- | --- | --- | --- |
| Fulton | 2561 | 4.13% | 5* | 5* | 25* |
| Glynn | 2193 | 0.71% | 3 | 5 | 15 |
| Camden | 1780 | 0% | 1 | 4 | 4 |
| Lowndes | 1430 | 0.58% | 3 | 4 | 12 |
| Ware | 911 | 0% | 1 | 3 | 3 |
| Baker | 605 | 0% | 1 | 3 | 3 |
| Dekalb | 431 | 6.67% | 5 | 2 | 10 |
| Emmanuel | 151 | 0% | 1 | 2 | 2 |
| Gwinnett | 149 | 1.61% | 4 | 1 | 4 |
| Clarke | 131 | 0% | 1 | 1 | 1 |
|  | 10,342 |  |  |  |  |

Though three additional counties, Glynn, Lowndes and Camdem, showed high *Anopheles* abundance with lower WNV transmission intensity, this suggests these counties could possibly emerge as potential hot-spots over a long term and possibly become vulnerable to malaria risk outbreaks in the nearest future (Figure 2 and Figure 3).

**Figure 3.**
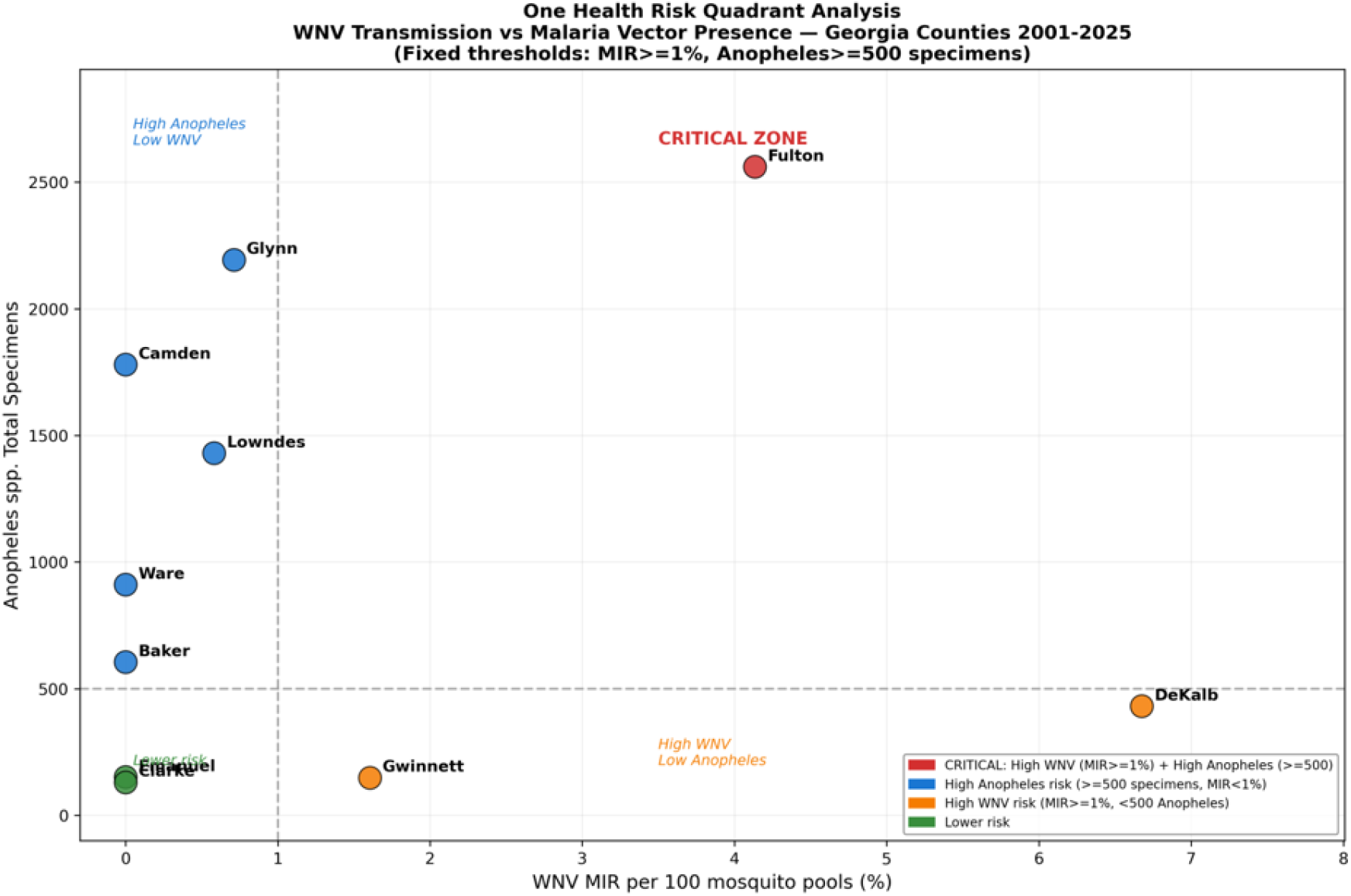
Risk quadrant analysis integrating WNV Minimum Infection Rate and *Anopheles* vector abundance by county in Georgia (2001–2025). Red = critical convergence zones (high WNV + high *Anopheles*); Blue = high *Anopheles* abundance, lower WNV; Orange = high WNV, low *Anopheles* abundance; Green = lower risk.

## DISCUSSION

The continuous presence and population dominance of *An. crucians, An. punctipennis*, and *An. quadrimaculatus* re-affirms the persistence of historically recognized malaria vector species within the state of Georgia. The dominance of these three species is consistent with their established epidemiological significance as competent malaria vectors across the United States. *An. quadrimaculatus* has historically been recognized as the principal malaria vector in the eastern United States, and together with *An. Punctipennis* which has been directly implicated in the 2002 Loudoun County, Virginia outbreak of *Plasmodium*-positive mosquitoes with confirmed human malaria cases since national surveillance began in 1957 (33,34,35-40).

This finding is consistent with the prior work of Dye-Braumuller and Kanyangarara (2021), who reported that Georgia had among the highest rates of county-level *Anopheles* reported nationally, with 89.31% of counties documenting at least one *Anopheles* species. The continued persistence of *Anopheles* species across Georgia’s landscape is consistent with recent surveillance findings from other southeastern and mid-Atlantic states such as Florida. In Miami-Dade County, Florida, *An. crucians* and *An. quadrimaculatus* were both recovered in substantial numbers from surveillance data collected between 2020 and 2022, with the anthropophilic feeding behavior of *An. quadrimaculatus*, and to a lesser extent *An. crucians*, which has been identified as a key factor underlying their continued relevance to malaria transmission risk in the United States (41).

Similarly, a 2026 analysis of *Anopheles* hot-spot in Maryland, encompassing all three species identified in the present study, found that low impervious surface percentage was the most influential predictor of *Anopheles* presence, followed by warmer, more humid micro-climates during the summer seasons which were also found to provide optimal conditions for Anopheles mosquito proliferation (42). These findings suggest that the environmental characteristics of Georgia’s coastal plain and Piedmont georegions may similarly support the sustained presence of these *Anopheles* species across the state. Beyond species-level patterns, the distribution of *Anopheles* abundance across Georgia’s counties revealed further insight into localized risk. Findings from this study indicate that significant *Anopheles* abundance was concentrated primarily within Fulton county which already has increased WNV MIR rates compared with lower severity in Gwinnett and Dekalb counties. This significant concentration of *Anopheles* populations within a limited number of counties, rather than an even distribution throughout the state, suggests that these counties may possess localized environmental and habitat conditions that disproportionately favor vector suitability.

However, *Anopheles* population abundance alone does not necessarily reflect overall malaria outbreak risk, as the presence of these competent vectors must coincide with active disease transmission to constitute a meaningful threat. For this reason, *Anopheles* abundance data were integrated with WNV transmission intensity (MIR) to identify counties representing convergent vector-borne disease risks. Among the counties assessed, Fulton county was the only county to meet the criteria for critical convergence status, exhibiting concurrently elevated WNV transmission intensity as well as the highest *Anopheles* population abundance. This convergence suggests that environmental or urban ecological drivers, such as dense human settlements, standing water associated with urban infrastructure, and localized micro-climatic conditions, may simultaneously favor both arboviral and malaria vector proliferation in Fulton County, positioning it as the highest-priority target for integrated vector surveillance in Georgia.

In contrast to the high vector abundance and high WNV intensity observed in Fulton county, other counties, including Glynn, Camden, Lowndes, and Ware, exhibited substantial *Anopheles* abundance despite comparatively low WNV transmission intensity. This pattern suggests that surveillance systems oriented solely toward arthropod-borne viral diseases such as WNV may fail to capture true malaria outbreak risk present in these counties. This highlights the need for the mosquito surveillance program to simultaneously track *Anopheles* mosquito populations, rather than only when WNV cases are already present. This is due to the fact that *Anopheles* mosquitoes were not detected in most of the same counties where WNV surveillance was conducted such as Chatham county which recorded zero *Anopheles* specimens, while counties with significant *Anopheles* populations (Camden, Ware, Glynn) had very low WNV activity. This geographic non-overlap further supports the ecological divergence between *Culex*-driven arboviral transmission and *Anopheles* associated malaria vector risk in Georgia.

Conversely, counties such as DeKalb and Gwinnett showed elevated WNV transmission intensity alongside comparatively low *Anopheles* abundance. This suggests that WNV transmission in these counties may be driven by other factors, such as the presence of *Culex* species, the principal WNV vector, as well as other localized ecological and urban conditions. This pattern demonstrates that WNV risk and malaria vector risk may not always driven by the same environmental conditions, thereby supporting the need to monitor multiple mosquito species concurrently. This multi-vector approach is consistent with the growing international application of One Health surveillance frameworks; for example, Gobbo et al. (2025) who demonstrated the integration of entomological, veterinary, and human surveillance data to monitor WNV and Usutu virus transmission in north-eastern Italy. In addition, vector abundance data have been shown to serve as a meaningful, though secondary, predictor of arboviral disease risk relative to demographic factors (44, 45), lending further support to its use as a component of integrated risk assessment in the present study.

Projections for the period 2025–2036 indicate that both *Anopheles* abundance and WNV transmission intensity are expected to increase gradually under current climate scenarios (Figure 4). Counties already identified as critical convergence zones, such as Fulton, may experience further intensification of dual vector-borne disease risk and possible outbreaks underscoring the importance of sustained, forward-looking surveillance and preparedness. These findings suggest that intensified *Anopheles* pool testing for circulating plasmodium species, especially *P. falciparum* and *P. vivax* may help support projections in malaria outbreak risk, rather than relying solely on current surveillance data. This will adequately ensure adequate preparedness for emerging transmission risk or outbreaks in high-priority counties of Georgia.

**Figure 4.**
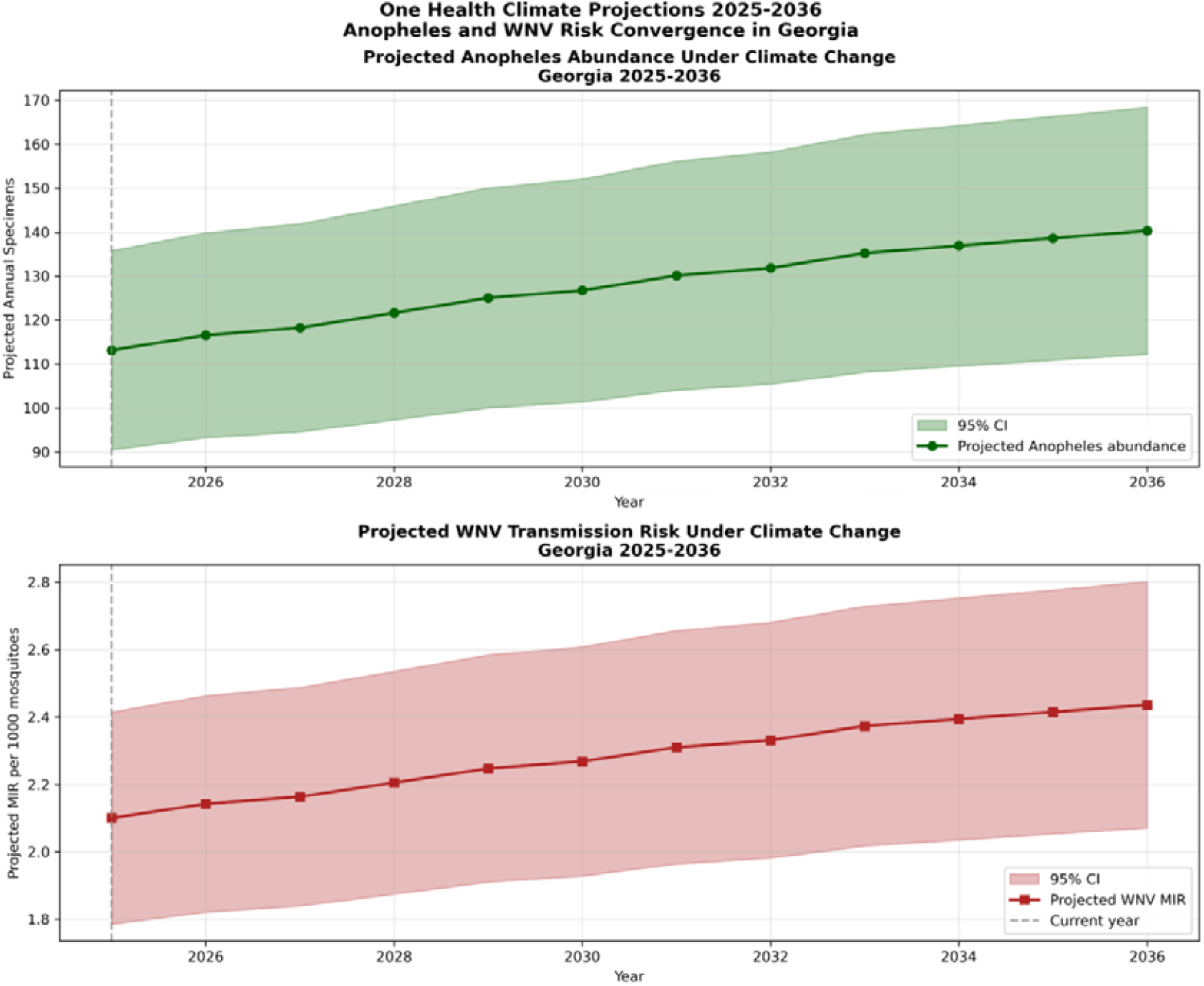
Projected trends in Anopheles abundance and WNV transmission risk in Georgia under moderate climate change scenarios (2025–2036). Panel A: Projected annual *Anopheles* abundance with 95% confidence interval. Panel B: Projected WNV Minimum Infection Rate with 95% confidence interval. The dashed line indicates the current year (2025).

## Conclusion

This statewide analysis of *Anopheles* distribution identified Fulton County as the principal locus of both convergent malaria and arboviral vector-borne disease risk in Georgia. Although autochthonous malaria has been absent from the United States for over seven decades, the persistence of competent *Anopheles* vector species across multiple Georgia counties, combined with rising international travel from endemic regions and projected climate-driven expansion of vector habitat suitability, necessitates continued mosquito surveillance and outbreak preparedness. Public health agencies in Georgia should prioritize integrated surveillance in identified high risk convergence zones but further establish baseline molecular surveillance to detect cryptic *Plasmodium* circulation.

## Data Availability

All data produced in the present study are available upon reasonable request to the authors

## Data Availability

Data sets are available upon request.

## Acknowlegdgement

The authors would like to appreciate the Georgian department of public health department and the Black fly research and resource center, University of Georgia.

## Notes

### Competing Interest Statement

The authors have declared no competing interest.

